# Occupational stress and quality of sleep among dentists in the Philippines

**DOI:** 10.1101/2021.03.23.21254135

**Authors:** Junhel Dalanon, Ivy Fernandez, Cora Estalani, Sachiko Chikahisa, Noriyuki Shimizu, Rozzano Locsin, Yoshizo Matsuka

## Abstract

**Background:** Occupational stress and sleep quality share a bidirectional relationship among other comorbidities. Dentistry remains among the health professions that suffer the greatest stress.

**Aim:** This study aimed to evaluate the correlation between occupational stress and sleep quality among dentists in the Philippines.

**Methods:** The Caregiver Self-Assessment Questionnaire assessed occupational stress and the Pittsburgh Sleep Quality Index measured sleep quality in purposively sampled dentists (n=420) in the Philippines.

**Results:** The prevalence of occupational stress (19%) was low yet poor sleep quality (71%) was high. The global PSQI score (p=0.047) and sleep onset latency (p=0.036) were found to be associated with occupational stress.

**Conclusions:** This study supports the vast literature on the relationship between occupational stress and sleep quality, despite documenting an unexpected low prevalence of occupational stress in Filipino dentists. Further research on the coping mechanisms of these dentists is warranted.

## INTRODUCTION

Occupational stress is naturally endemic across all disciplines, but abnormally high levels and chronicity can have a detrimental effect on a person (1-3). The perception and outlook of a patient towards a disease condition could affect the stress level of a healthcare professional (4). The presence of confounding variables or collective mental health states such as depression, anxiety, and sleep problems have been found to be positively correlated to occupational stress (5).

Globally, dentistry is one of the most stressful among the health professions due to the working conditions and the nature of work. Recently, it was found out that more than half (54.9%) of the dentists of the British Dental Association experience a high degree of stress. Almost half of these dentists could not cope with the level of stress in their job and almost a fifth (17.6%) confessed of seriously contemplating on committing suicide. The stress levels were high across all dental specializations but highest in general practitioners. Complaints or litigations (79%) is the most common source of stress (6). The elevation of stress in the practice of dentistry can be due to the maintenance of posture over a long period of time, visual difficulty due to the small size of the oral cavity, noise and odor in the work environment, and the persistent desire for technical perfection (3, 7-10).

Even though there are several articles chronicling the relationship between work stress and sleep in dentists, there is an absence of information regarding their existence in Filipino dentists. A substantial number of articles on occupational stress were done on Filipino migrant nurses, but only a few were made in the Philippines (11). The dental caries prevalence in the country is among the highest in the world, the willingness-to-pay for dental services very low, and their personalities do not fit that of a dentist (12-13). These are potential stressors to the local dentists. The purpose of this study was to examine the sleep quality and occupational stress of the dentists in the Philippines through self-reported measures. Moreover, this study aimed to uncover the confounding correlation between stress and sleep with other socio-demographic variables among dentists in the Philippines.

## METHODS

Data collection. This cross-sectional study and all other related procedures were done in accordance with the Declaration of Helsinki (14) and the research guidelines of the School of Dentistry of Southwestern University PHINMA. Ethical approval was sought and approved from the latter. Integrated self-reported questionnaires were given to 420 dentists, who are members of the Philippine Dental Association (PDA). The participants were informed that they were conveniently sampled, their responses will be kept confidential, their identities will remain anonymous, and participation is completely voluntary. Informed consent were sought from the participants before they were allowed to continue with the survey. The socio-demographic information gathered include age, gender, type of practice, and the presence of back pain.

Sleep quality measures. Supplementing the socio-demographic data, sleep quality was assessed using the Pittsburgh Sleep Quality Index (PSQI) developed by Buysse et al. (15). The PSQI was used to gauge the quality of sleep and pattern for a specified month. It is a validated clinical and non-clinical measure for assessing seven factors of sleep quality including subjective sleep quality, sleep latency, sleep duration, habitual sleep efficiency, sleep disturbances, use of sleeping medications, and daytime dysfunction. It uses a 4-point Likert scale (0-3) where the scores from each component are summed to generate a global score (0-21). A global PSQI score greater than 5 indicates poor sleep quality. All-in-all, this is a research tool that can give an effective and inexpensive assessment of the sleep quality of the participants.

Occupational stress measures. Originally developed and published by the American Medical Association (16), the Caregiver Self-Assessment Questionnaire (CSAQ) measures work-related or occupational stress. It is unique as the questions are directed towards the healthcare provider and patient relationship. The CSAQ had also been used in the past as a pre-screening measure for depressive symptoms. This valid and easy-to-use self-report measure is an 18-item questionnaire where the first 16 questions are answerable with a dichotomous “Yes” or “No”. The last two questions are to be answered with a scale of 1 to 10. A high degree of stress is assumed if the participant answered “Yes” to either or both questions 4 (feeling of being overwhelmed) and 11 (crying spells), the total “Yes” score is 10 or more, and question 17 (perception of stress) and 18 (perception of health) is rated 6 or more.

Data analysis. Data were tallied and analyzed with the use of a statistical software package (SPSS 21). The socio-demographic profile of the participants and the PSQI factors were presented using descriptive statistics. Continuous variables were presented by mean and standard deviation, while categorical variables were organized in frequency and percentages. The association between occupational stress as a dependent variable with the socio-demographic profile and sleep quality was calculated using Pearson’s Chi-square Test.

## RESULTS AND DISCUSSION

The descriptive data in Table 1 shows that majority of the respondents were at the age range of 21-30 years old (n=230, 54.8%), female (n=350, 83.3%), engaged in private practice (n=280, 66.7%), and suffering from back pain (n=370, 88.1%). The table also shows that most of the participants are unexpectedly not stressed (n=340, 81.0%) yet are suffering from poor sleep quality (n=300, 71.0%).

**Table 1.**
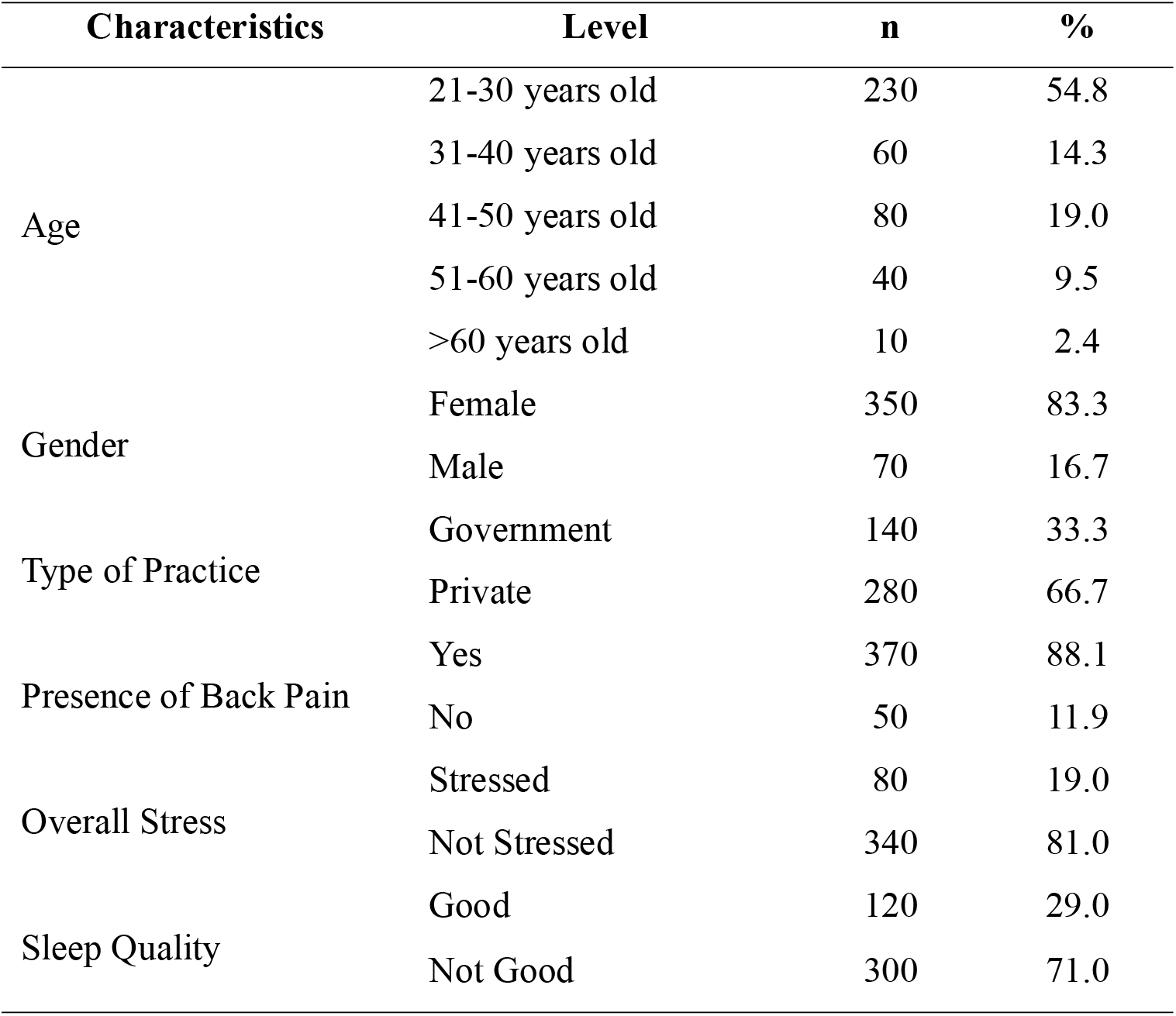
Socio-demographic profile of the dentists in the Philippines (n=420)

| **Characteristics** | **Level** | **n** | **%** |
| --- | --- | --- | --- |
| Age | 21-30 years old | 230 | 54.8 |
|  | 31-40 years old | 60 | 14.3 |
|  | 41-50 years old | 80 | 19.0 |
|  | 51-60 years old | 40 | 9.5 |
|  | >60 years old | 10 | 2.4 |
| Gender | Female | 350 | 83.3 |
|  | Male | 70 | 16.7 |
| Type of Practice | Government | 140 | 33.3 |
|  | Private | 280 | 66.7 |
| Presence of Back Pain | Yes | 370 | 88.1 |
|  | No | 50 | 11.9 |
| Overall Stress | Stressed | 80 | 19.0 |
|  | Not Stressed | 340 | 81.0 |
| Sleep Quality | Good | 120 | 29.0 |
|  | Not Good | 300 | 71.0 |

The global PSQI score of the Filipino dentists is high (x□=6.88, s=3.33), although subjective sleep quality (x□=1.02, s=0.64), sleep latency (x□=1.52, s=0.94), sleep duration (x□=1.05, s=0.94), habitual sleep efficiency (x□=0.88, s=1.13), sleep disturbances (x□=1.17, s=0.54), use of sleeping medication (x□=0.21, s=0.61), and daytime dysfunction (x□=1.02, x□=0.64) remain good (Table 2).

**Table 2.** Global PSQI Score and the factors of sleep quality (n=420)

| **Characteristics** | **Mean** | **SD** |
| --- | --- | --- |
| Global PSQI Score | 6.88 | 3.33 |
| Subjective Sleep Quality | 1.02 | 0.64 |
| Sleep Latency | 1.52 | 0.94 |
| Sleep Duration | 1.05 | 0.94 |
| Habitual Sleep Efficiency | 0.88 | 1.13 |
| Sleep Disturbances | 1.17 | 0.54 |
| Use of Sleeping Medication | 0.21 | 0.61 |
| Daytime Dysfunction | 1.02 | 0.64 |

PSQI – Pittsburgh Sleep Quality Index, SD – Standard Deviation

**Table 3.**
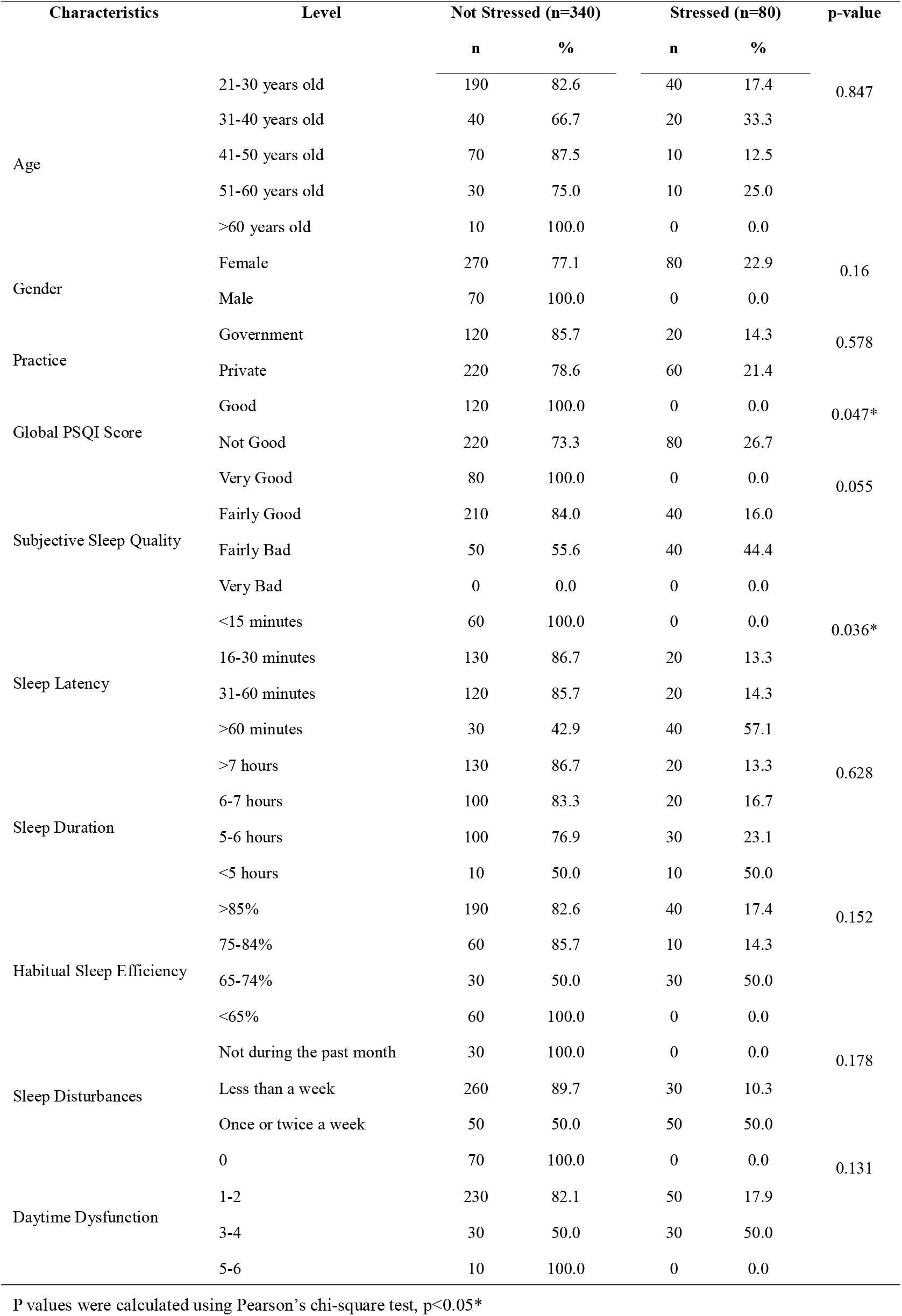
Correlational analysis between occupational stress with the socio-demographic profile and sleep quality.

The cohorts with the highest percentage for the presence of stress are 31-40 years old (n=20, 33.3%), female (n=80, 22.9%), private practitioners (n=60, 21.4%), Global PSQI >5 (n=80, 26.7%), fairly bad subjective sleep quality (n=40, 44.4%), sleep latency >60 minutes (n=40 57.1%), sleep duration of <5 hours (n=10, 50.0%), habitual sleep efficiency of 65-74% (n=30, 50.0%), those experiencing sleep disturbances once or twice a week (n=50, 50.0%), and those that scored 3-4 for daytime dysfunction (n=30, 50.0%). The global PSQI score (p=0.047) and sleep onset latency (p=0.036) were found to be associated with occupational stress.

Presently and to the knowledge of the researchers, this is the first ever study that has gauged the occupational stress and sleep quality of dentists in the Philippines. Although a literature review and meta-analyses showed that blue collared workers were the most stressed working group, dentists remain among the most stressed of the health professionals (17). In a developing country like the Philippines, biochemical tests like salivary cortisol for stress (18) or biophysical assays like polysomnography for sleep quality (19) are impractical and costly. The validated and easy-to-use self-reported questionnaire like the CSAQ and PSQI are more appropriate. High caries prevalence, personalities not suitable for being a dentist, patients’ unwillingness to pay for dental services, and the healthcare providers’ perception of the patients’ condition, these are only some of the source of stressors of the dental professionals in the Philippines (12-13). Gordon (4) elaborated on shared vulnerability, or a healthcare provider’s perception of a patient’s stigmatizing condition, as a potential source of stressor. The CSAQ is a distinctive measure of stress as it relates, not only to the physical factors of work, but also the psychosocial aspect. The presence of back pain is also the same with the Global Burden of Disease Study, which is also a potential source of stressor.

A distinct finding of this study is the low prevalence of stress in dentists. Contrary to other findings, dentistry is one of the most stressful disciplines in healthcare. In the Philippine setting, de Castro found that acute and chronic effects of stress and overwork was a top-ranking concern in nurses in the Philippines. Although this study is incomparable to the as the dentists’ and nurses’ nature of work differ considerably. This study was also limited to the CSAQ in terms of self-reported stress in relation to the healthcare professionals’ view or concern for the patient (16).

In this study, occupational stress was not found to be associated with either age, gender, or type of practice. In addition, neither were subjective sleep quality, sleep duration, habitual sleep efficiency, sleep disturbances, and daytime dysfunction related to the stress of the Filipino dentists. Meanwhile, a PSQI score of 6.88 ± 3.33 and 71% of the participants in this study were consistent with the findings in a report on Korean dentists (5). Augner (20) in Austrian nursing students, Hayashino et al. (21) in Japanese dental population, Kaneita et al. (22) in Japanese high school students, and Song et al. (5) in Korean dentists found correlations between sleep and mental health states. Currently, this study shows that stress, which is part of the mental health states, is correlated with the global PSQI score and sleep onset latency.

There are about tens of thousands of dentists in the Philippines that are unaccounted for and this is one of the limitations of the study. As dentistry is considered a profession in the Philippines which is predominantly for females, the high turnout of female participants (83.3%) is another limitation. Despite this fact, this study has given ample information on the occupational stress and sleep quality of dentists in the Philippines. Since the low prevalence of occupational stress is a unique finding in dentists, further studies should be done aimed at the sources of stress and coping mechanisms of the dentists in the Philippines. Comparisons can also be drawn out between dentists, dental hygienists, dental technologists, and dental students.

## CONCLUSION

The results of this study determined that majority of dentists in the Philippines do not experience stress but perceived that they have poor sleep quality. Furthermore, this also demonstrated that the global PSQI score and sleep onset latency are associated with occupational stress. The recommendation of PSQI and CSAQ as a cost-effective and invaluable tool in measuring sleep quality and occupational stress in dentists of developing countries should be considered.

## Data Availability

Data from the authors is available upon request.

## Acknowledgments

This research study was funded in part through the International Association for Human Caring Shirley C. Gordon Award for advancing the theory of Shared Vulnerability.

## REFERENCES

1. Wenjuan WA, Hui RE, Qiuye TI, Chunling TA, Wenjuan ME. Effects of Occupational Stress on Blood Lipids, Blood Sugar and Immune Function of Doctors. Iranian Journal of Public Health. 2019 May 14;48(5):825–33.

2. Rada RE, Johnson-Leong C. Stress, burnout, anxiety and depression among dentists. The Journal of the American Dental Association. 2004 Jun 1;135(6):788–94.

3. Sancho FM, Ruiz CN. Risk of suicide amongst dentists: myth or reality?. International dental journal. 2010 Dec;60(6):411–8.

4. Gordon SC. Shared vulnerability: a theory of caring for children with persistent head lice. The Journal of School Nursing. 2007 Oct;23(5):283–92.

5. Song KW, Choi WS, Jee HJ, Yuh CS, Kim YK, Kim L, Lee HJ, Cho CH. Correlation of occupational stress with depression, anxiety, and sleep in Korean dentists: cross-sectional study. BMC psychiatry. 2017 Dec;17(1):398.

6. Collin V, Toon M, O’Selmo E, Reynolds L, Whitehead P. A survey of stress, burnout and well-being in UK dentists. British Dental Journal. 2019 Jan;226(1):40–49.

7. Alexander RE. Stress-related suicide by dentists and other health care workers: Fact or folklore?. The Journal of the American Dental Association. 2001 Jun 1;132(6):786–94.

8. Möller AT, Spangenberg JJ. Stress and coping amongst South African dentists in private practice. The Journal of the Dental Association of South Africa= Die Tydskrif van die Tandheelkundige Vereniging van Suid-Afrika. 1996 Jun;51(6):347–57.

9. Lang-Runtz H. Stress in dentistry: it can kill you. Journal (Canadian Dental Association). 1984 Jul;50(7):539–41.

10. Kang YS, Kam S, Lee SW, Chun BY, Yeh MH. Job Stress and Its Related Factors in South Korean Doctors. Journal of Preventive Medicine and Public Health. 2001 May 1;34(2):141–8.

11. De Castro AB, Cabrera SL, Gee GC, Fujishiro K, Tagalog EA. Occupational health and safety issues among nurses in the Philippines. Aaohn Journal. 2009 Apr;57(4):149–57.

12. Dalanon J, Diano LM, Esguerra R, Belarmino MP, Docor MR, Rodis OM, Locsin R, Matsuka Y. The Cebuano Mothers’ Willingness to Pay for Dental Healthcare. The Journal of the Philippine Dental Association. 2018 Dec;65(2):33–37.

13. Dalanon J, Matsuka Y. An Investigation of the Filipino Dental Students’ Personality Types. Philippine Journal of Health Research and Development. 2018 Nov 14;22(2):12–7.

14. World Medical Association declaration of Helsinki. Recommendations guiding physicians in biomedical research involving human subjects. JAMA. 1997 Mar 19;277(11):925–6.

15. Buysse DJ, Reynolds III CF, Monk TH, Berman SR, Kupfer DJ. The Pittsburgh Sleep Quality Index: a new instrument for psychiatric practice and research. Psychiatry research. 1989 May 1;28(2):193–213.

16. EpsteinLLubow G, Gaudiano BA, Hinckley M, Salloway S, Miller IW. Evidence for the validity of the American medical association’s caregiver selfLassessment questionnaire as a screening measure for depression. Journal of the American Geriatrics Society. 2010 Feb;58(2):387–8.

17. Milner A, Spittal MJ, Pirkis J, LaMontagne AD. Suicide by occupation: systematic review and meta-analysis. The British Journal of Psychiatry. 2013 Dec;203(6):409-|

18. Šuşoliaková O, Šmejkalová J, Bičíková M, Hodačová L,Málková A, Fiala Z. Assessment of Work-Related Stress by Using Salivary Cortisol Level Examination among Early Morning Shift Workers. Central European journal of public health. 2018 Jun 1;26(2):92–7.

19. Smardz J, Martynowicz H, Wojakowska A, Michalek-Zrabkowska M, Mazur G, Wieckiewicz M. Correlation between Sleep Bruxism, Stress, and Depression—A Polysomnographic Study. Journal of clinical medicine. 2019 Sep;8(9):1344.

20. Augner C. Associations of subjective sleep quality with depression score, anxiety, physical symptoms and sleep onset latency in students. Central European journal of public health. 2011 Jun 1;19(2):115.

21. Hayashino Y, Yamazaki S, Takegami M, Nakayama T, Sokejima S, Fukuhara S. Association between number of comorbid conditions, depression, and sleep quality using the Pittsburgh Sleep Quality Index: results from a population-based survey. Sleep Med. 2010 Apr;11(4):366–71.

22. Kaneita Y, Yokoyama E, Harano S, Tamaki T, Suzuki H, Munezawa T, et al. Associations between sleep disturbance and mental health status: a longitudinal study of Japanese junior high school students. Sleep Med. 2009 Aug;10(7):780–6.

